# A 3D approach to understanding heterogeneity in early developing autisms

**DOI:** 10.1101/2024.05.08.24307039

**Authors:** Veronica Mandelli, Ines Severino, Lisa Eyler, Karen Pierce, Eric Courchesne, Michael V. Lombardo

## Abstract

Phenotypic heterogeneity in early language, intellectual, motor, and adaptive functioning (LIMA) features are amongst the most striking features that distinguish different types of autistic individuals. Yet the current diagnostic criteria uses a single label of autism and implicitly emphasizes what individuals have in common as core social-communicative and restricted repetitive behavior difficulties. Subtype labels based on the non-core LIMA features may help to more meaningfully distinguish types of autisms with differing developmental paths and differential underlying biology. Using relatively large (n=615) publicly available data from early developing (24-68 months) standardized clinical tests tapping LIMA features, we show that stability-based relative cluster validation analysis can identify two robust and replicable clusters in the autism population with high levels of generalization accuracy (98%). These clusters can be described as Type I versus Type II autisms differentiated by relatively high versus low scores on LIMA features. These two types of autisms are also distinguished by different developmental trajectories over the first decade of life. Finally, these two types of autisms reveal striking differences in functional and structural neuroimaging phenotypes and their relationships with gene expression. This work emphasizes the potential importance of stratifying autism by a Type I versus Type II distinction focused on LIMA features and which may be of high prognostic and biological significance.

---

The diagnostic concept of autism has been anything but stationary over time^1^. To illustrate this non-stationarity, consider that the autism of several decades ago was a population largely comprised of individuals with the core social-communication (SC) and restricted repetitive behavior (RRB) issues, but also frequently possessed profound issues in non-core features such as language, intellectual, motor, and adaptive functioning (features that are henceforth referred to as acronym ‘LIMA’). In contrast, the autism of today is largely comprised of individuals with varying and sometimes subtle core SC and RRB issues, but whom are much more likely to not have such profound LIMA difficulties^2^. As a likely result of these changes over time, we have seen phenomena such as increasing heterogeneity^3^, prevalence^4^, decreasing effect sizes^5^, and a change in views about autism, particularly from the neurodiversity perspective. Thus, it is no surprise that the most contentious debates in the field today deal with topics that ultimately boil back down to contrasting the relative importance of core SC and RRB versus non-core LIMA features. This can be seen when in debates contrasting medical versus social perspectives^6^, vocabulary and language use^7^, and debate regarding different subtyping approaches (e.g., prototypical autism, profound autism)^8–10^ versus ‘spectrum’/continuous models (e.g., autistic traits, polygenic risk)^11,12^.

Our contention is that the autism spectrum has become too wide to be meaningful as a whole when it comes to finding explanations about differential ***b***iology, ***o***utcomes, and responses to ***t***reatment (i.e. an acronym we label as the ‘BOT’ objectives^1^). Thus, at least for the purposes of the BOT objectives, we suggest that autism must be split into different types of autisms distinguished by non-core LIMA features. Owing to terminology already in use in the literature regarding dual perspectives of autism as ‘disability’ (i.e. the medical model) versus a ‘difference’ (i.e. social models such as neurodiversity), we propose that an initial split of autism be one of two highly differentiated early developmental and behavioral types – that is, one of profound ‘disability’ (Type I) versus one of a ‘difference’ (Type II). Furthermore, these contrasting types are hypothesized to be most prominently differentiated over ‘development’. Putting together these 3 ‘D’ words – ‘disability’ versus ‘difference’ over ‘development’ – we propose a framework called AUTISMS-3D (henceforth referred to as A3D). In simple terms, the A3D model suggests that when one focuses on the non-core LIMA features, autism can be thought of as at least two different types that contrast disability versus difference over development.

A key distinction separating A3D from other approaches^8,9,13^ is the idea that these theorized autism subtypes are evident and detectable with unsupervised data-driven approaches in early development. In past work utilizing just the domains of the Vineland Adaptive Behavior Scales (VABS), we showed that stability-based relative clustering validation could be applied to separate the predicted Type I versus Type II autisms with very high generalizability accuracy in new unseen datasets^14^. Thus, unsupervised data-driven discoveries can be immediately translated into supervised prediction models that can generalize with very high accuracy and highlight the A3D subtype distinction. This attribute of the A3D approach separates it from past issues that have been present with subtyping in autism. One of the primary reasons for abandoning DSM-IV subtype labels of PDD-NOS, Asperger Syndrome, high- and low-functioning autism labels was because of the lack of agreement between expert clinicians in making such labels^15^. Even with more recent suggestions such as the label of ‘profound autism’^9^, it could be argued that the defining boundaries for that label are still based on potentially arbitrary and/or hard cutoffs for age (e.g., >8 years) and IQ (e.g., IQ<50). Furthermore, the Lancet commission suggested the use of a label like profound autism be primarily administrative^9^, leaving a gap with regard to how research might be able to utilize a label that picks up on a similar kind of conceptual distinction. In contrast to the approach of focusing on ‘prototypical autism’, A3D again separates itself by not relying on subjective expert clinical judgement for making such a distinction. While there is no substitute for expert clinical judgement as the gold standard for making the autism diagnosis itself, for making these types of subtype distinctions A3D makes the prediction that subtypes based on LIMA features will be evident in robust statistical differences that unsupervised data-driven models will be able to detect with very high generalizability accuracy. In this work, we go beyond past work utilizing VABS domains^14^ to measure early developing skills in all four pillars of non-core LIMA features. We predict that Type I versus Type II autisms will be evident via stability-based relative clustering validation^16^. Furthermore, we predict that the theorized Type I versus Type II autisms will be heavily differentiated in developmental trajectories tested with longitudinal data over the first 5-6 years of life.

Given one of the field’s over-arching research priorities is to better understand differential biology characteristic of many autisms (i.e. the ‘B’ in the ‘BOT’ objectives)^1^, another goal of the A3D model is to test hypotheses regarding whether these two phenotypically differentiated types are also sensitive indicators of differential underlying neurobiology. In related past work, we utilized a stratification approach based on early language outcomes (ELO) and showed that the distinction between good versus poor ELO is indicative of types with differential underlying early neurobiology^17–20^. However, this stratification approach leverages only one of the four pillars of non-core LIMA features we hypothesize to be relevant (i.e. language). Thus, in the current work, we seek to evaluate how autism subtypes predicted from A3D (based on the full set of LIMA features) may match up in terms of early functional neural systems response to speech and structural neuroimaging phenotypes (e.g., surface area and cortical thickness) that had been previously identified to be highly important in ELO subtypes^17,18^.

## Methods

### NDA dataset

The primary dataset used in this work for the purposes of building a generalizable stratification model comes from the National Institute of Mental Health Data Archive (NDA) (https://nda.nih.gov/). NDA includes several datasets that measure two standardized early clinical measures that are integral for this work – the Mullen Scales of Early Learning (MSEL) and Vineland Adaptive Behavior Scales (VABS). All MSEL and VABS data from NDA was downloaded in February 2022 for further analysis in this work. MSEL and VABS data from NDA was then filtered, preprocessed and analyzed for the existence of stable and highly replicable theoretically predicted autism subtypes (e.g., Type I and Type II) in early ages via unsupervised data-driven approaches (i.e., *reval*)^16^. We downloaded all available MSEL^21^ and VABS (i.e., VABS 2nd edition parent and caregiver form or survey form; VABS 3rd edition)^22,23^ within the age range of 0-68 months. VABS standard scores from communication, daily living skills, socialization, and motor domains were extracted for use in further data analysis. For MSEL data, we extracted chronological age at MSEL assessment as well as age-equivalent scores for visual reception (VR), fine motor (FM) and receptive and expressive language (RL; EL) subscales. Next, we implemented a series of data cleaning steps that entailed selecting only individuals with an autism diagnosis, dropping duplicate data, extracting the earliest available time point for individuals with longitudinal data, selecting only individuals whose MSEL and VABS data were assessed not more than one year apart, and finally drop individuals with more than one missing subscale for each of the instruments used (Fig 1A). After data cleaning, a total of n=615 autistic individuals remained for further downstream analysis. Data from individuals with more than one data point were also utilized for the subsequent longitudinal analysis sought to test the hypothesis that the subtypes’ developmental trajectories heavily differentiate over the first 5-6 years of life (MSEL n=179; VABS n=354).

**Figure 1:**
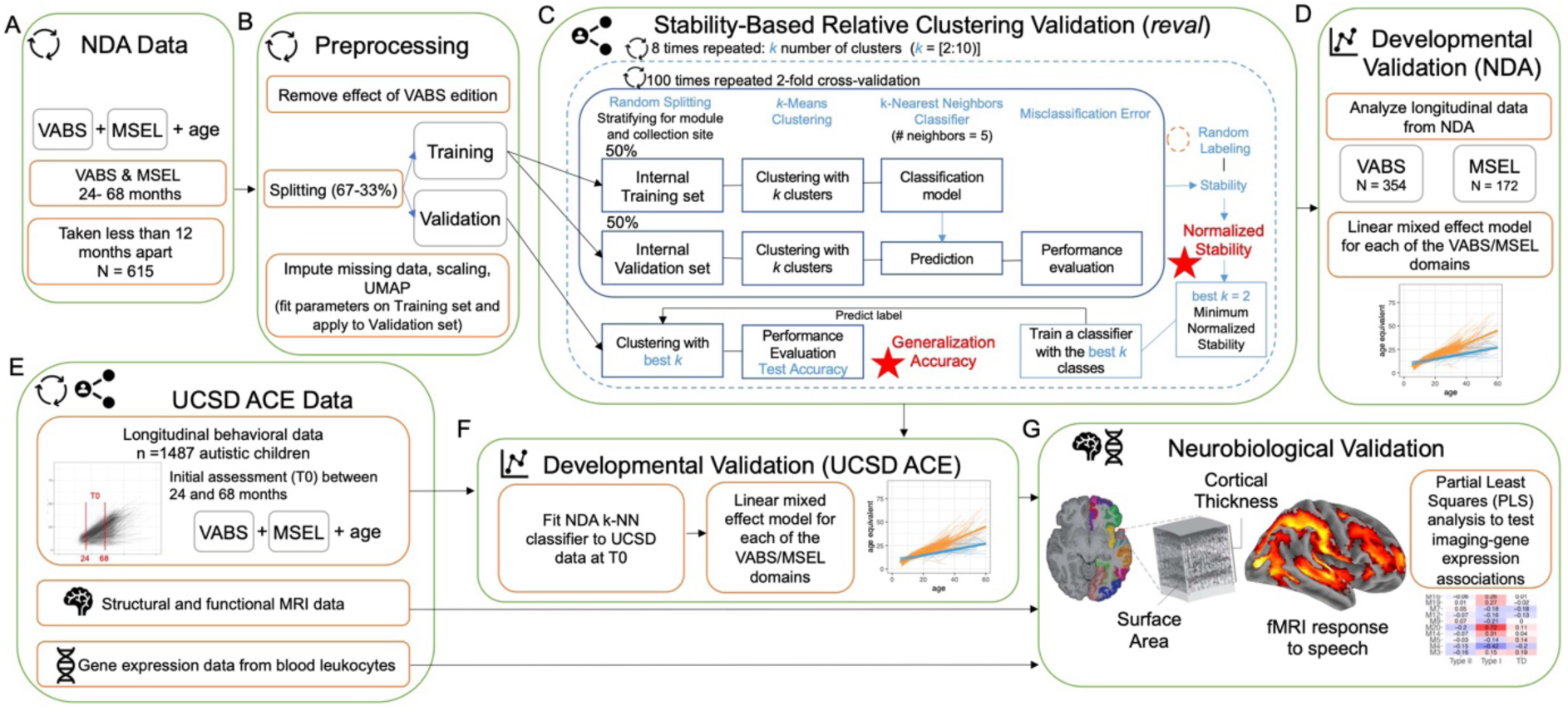
Schematic of data analysis workflow/pipeline. This figure shows a workflow of the analyses conducted in this work. Panels A and B depict how we select, clean, and preprocess data from the National Institute of Mental Health Data Archive (NDA). To identify subtypes in a data-driven manner, we used stability-based relative clustering validation analysis (reval) (panel C) on a final sample of 615 autistic children aged 24-68 months. Subtypes are tested for their prognostic validity (panel D) utilizing longitudinal data from NDA. The stratification model was tested again for developmental trajectory differences in an independent longitudinal dataset from UCSD ACE (panel E-F). Subtypes were also tested for neurobiological differentiation using structural and functional neuroimaging data and relationships to blood leukocyte gene expression patterns (panel G).

Before doing the primary downstream *reval* clustering analysis on the NDA dataset, we had to address the batch effect caused by the presence of different VABS versions (e.g., VABS Edition 2 which comprises either the caregiver or survey forms, as well as VABS 3rd edition). For each of the VABS domains, we apply a batch correction factor to adjust the raw data. These factors correspond to the beta coefficients estimated in a linear model with VABS standard scores as the dependent variable and VABS version as the independent variable with age and sex as covariates). Batch correction factors are taken from Mandelli et al.,^14^ since they were estimated in a bigger sample size within the age range of interest for this work (n= 604, age 0-72 months). To project out batch-related variability brought on by the VABS version, the correction factors (i.e., beta coefficients) were multiplied against the raw data and then the difference was removed from the raw VABS domain standard scores. Subsequent *reval* clustering analyses were carried out with version-corrected VABS data.

### UCSD dataset

To test the stratification model for differences in developmental trajectories using a dataset that is independent of the NDA dataset used to build the stratification model, we utilized a longitudinal phenotypic dataset from the University of California, San Diego (henceforth referred to as UCSD ACE). This study was approved by the Institutional Review Board at University of California, San Diego. This dataset comprises longitudinal VABS and MSEL measurements from n = 1487 autistic children. The enrollment of most individuals into the UCSD ACE dataset started from ∼12 months using a combination of population-based autism risk screening and community referrals ^24^. Longitudinal data was collected every six months from the initial intake assessment until the age of 5-6 years of age, and included a comprehensive battery of assessments such as VABS, MSEL, and the Autism Diagnostic Observation Schedule (ADOS-2)^25^. Clinical diagnosis of autism was made at by expert clinicians at the UCSD ACE center. To match the age range of the NDA subtyping model, we only included a subset of children with an assessment between the ages of 24 and 68 months in the current study. The NDA subtype prediction model was used to predict subtype labels for the UCSD ACE dataset based on VABS -MSEL scores at the first assessment time point between 24-68 months.

A subset of autistic and non-autistic from the UCSD dataset (autism n=81; n=37 typically developing (TD); n=19 language or developmental delays (LD/DD)) with structural (MRI) and functional MRI (fMRI) and blood leukocyte gene expression data was also analyzed. This dataset was previously reported in past papers examining heterogeneity in early language outcomes (ELO)^17–19^. Here we applied the A3D prediction model to label autistic individuals in this imaging dataset as either Type I and Type II and then test if such labels are sensitive to differential underlying neurobiology. For details on data collection and processing of UCSD neuroimaging and gene expression data, see the section on neuroimaging and gene expression data.

### Phenotypic measures utilized in NDA and UCSD ACE datasets

#### Vineland Scales of Adaptive Behavior (VABS)

The VABS is a widely used standardized and semi-structured caregiver interview for assessing adaptive functioning in typical and clinical populations throughout the lifespan^22,23^. For children within the age range included in this work (≤ 68 months) the VABS assesses adaptive functioning skills within four domains: communication, daily living skills, socialization, and motor skills. In this work, we used standardized scores for each domain. Standardized scores are computed to indicate where an individual scores relative to typically developing age-appropriate norms, whereby for each standardized score, the mean is 100 and the standard deviation is 15.

#### Mullen Scales of Early Learning (MSEL)

The MSEL is a standardized assessment for non-verbal cognitive, motor, and language skills that can be administered in children aged 0-68 months^21^. In this work, four MSEL subscales were utilized which are relevant for assessing non-core autism characteristics within the A3D framework: visual reception (VR), fine motor (FM), receptive language (RL), and expressive language (EL). For each subscale, Developmental quotients (DQ) were computed as (age-equivalent/chronological age)*100, to have age-standardized scores centered around a value of 100. Computation of MSEL DQ scores eliminates some known issues with floored MSEL t-scores but also allows for MSEL to be used in further data analysis for clustering with compatible age-standardized scores consistent with VABS (e.g., age-standardized scores centered at a value of 100 to indicate normatively age-appropriate skill level). For evaluating MSEL developmental trajectories, we utilized MSEL age-equivalent scores.

#### Stability-based relative clustering validation analysis (reval)

Our primary stratification model utilizes an unsupervised data-driven clustering approach that seeks to identify the optimal number of clusters with optimal level of generalizability in new data. This approach is based on stability-based relative clustering validation^16,26^, whereby we transform the unsupervised learning framework into a classification problem which allows for immediate translation of data-driven clustering solutions into supervised classification models that can be empirically demonstrated to be robust and generalizable partitions of the autism population. Stability-based relative clustering validation analysis was implemented with our Python library called *reval*^16^. The *reval* algorithm is shown in Fig 1C and can be described in the following series of steps. First, the dataset is split into independent training (dataset ***X_tr_***) and validation (dataset ***X_val_***) sets. Within the training set (***X_tr_***), *reval* splits the data into an internal training (***X_tr_tr_***) and test (***X_tr_ts_***), with the goal to identify the optimal number of clusters (***k***). Clustering algorithm ***Α_k_*** is then fit to both ***X_tr_tr_*** and ***X_tr_ts_*** and produces clustering labels ***Y_tr_tr_*** and ***Y _tr_ts_*** respectively. A classifier (***Φ_tr_tr_***) is then trained on [***X_tr_tr_****, **Y******_tr_tr_***] and then fit to ***X_tr_ts_*** to predict labels ***Y_tr_ts_pred_***. Misclassification error is computed by comparing the ***Α_k_***’s clustering labels (***Y_tr_ts_***) to a classifier’s (***Φ_tr_tr_***) predicted labels (***Y_tr_ts_pred_***). The *reval* algorithm iterates this procedure over a range of possible ***k*** clusters, and then identifies the optimal ***k*** that minimizes misclassification error on the internal test set (***X_tr_ts_***). Misclassification error is the normalized Hamming distance between the actual clustering labels (***Y_tr_ts_***) versus the classifier’s (***Φ_tr_tr_***) predicted labels (***Y_tr_ts_pred_***). However, because of the non-uniqueness of clustering labels, *reval* permutes the labels (***Y_tr_ts_***) and uses the Kuhn-Munkres algorithm to identify the labeling solution that minimizes misclassification error. This final measure of misclassification error is called ‘clustering stability’^26^ and this index ranges from 0-1, with lower values indicating more stable and reproducible clustering solutions. Stability is then normalized to the stability of random labelings to arrive at the final measure of normalized stability. Thus, the overall first goal in *reval* is to identify the clustering solution ***Α_k_*** that minimizes normalized stability. Once the optimal ***k*** is identified from the training set, we then train a classifier (***Φ_tr_***) on the entire training set at the optimal ***k***, and then apply it to held-out validation set (***X_val_***). Clustering labels (***Y_val_***) obtained on the validation set ***X_val_*** are then compared to the classifier’s (***Φ_tr_***) predicted labels (***Y_val_pred_***) and we compute a measure of generalizability accuracy operationalized as 1-misclassification error. Clustering solutions with high generalizability accuracy are the solutions we seek, as they are informative to solutions that have high potential to generalize well on the larger autism population. Furthermore, if generalization accuracy is very high, the output of *reval* is a classifier (i.e. prediction model) which we can use on new independent datasets to predict subtype labels (e.g., Type I vs Type II).

Before running *reval* we first select data and features from the NDA dataset for use in clustering. For selecting NDA data, we utilized all available data from participants that had both MSEL and VABS between 24-68 months, and whereby the MSEL and VABS are sampled within 12 months of each other. For individuals with repeated MSEL and VABS measures (i.e. longitudinal data) the very first snapshot of MSEL and VABS was utilized for clustering. The features selected comprise all MSEL subscales (EL, RL, VR, FM) and VABS domains (Socialization, Communication, Daily Living Skills, Motor). Standardized scores from both measures were utilized for clustering, alongside the age at time of assessment. VABS standardized scores were already available on NDA. However, for the MSEL subscale scores are represented on NDA as age-equivalent scores.

Before entering the *reval* clustering analysis, the data is split into independent training (***X_tr_***) and validation (***X_val_***) sets (training set = 67%; test set = 33%) while balancing the splits for variables such as age, sex, and VABS version. Three preprocessing steps are then applied. First, we impute missing values with a k-Nearest Neighbors imputation algorithm where the number of neighbors is identified via grid search (*sklearn.impute.KNNImputer*). The next two steps are ones where the parameters are fit on the training set and then applied to the validation set - scaling each feature in the dataset to mean of 0, standard deviation (SD) of 1 (*sklearn.preprocessing.StandardScaler)* and applying the dimensionality reduction technique Uniform Manifold Approximation and Projection (UMAP)^27^ (https://umap-learn.readthedocs.io/en/latest) (n_neighbors = 30, min_dist = 0.0, n_components = 2, random_state = 42, metric = Euclidean) to reduce the number of features from 8 to 2 (Fig 1B).

While the *reval* clustering approach is meant to ensure the stability of clustering solutions, the optimal ***k*** that is determined must still be tested for whether it is a solution indicative of a situation where actual true clusters exist. To test the solution against the null hypothesis that no clusters are apparent in the data, we utilized a Monte Carlo simulation framework meant to test whether the data originate from a single multivariate Gaussian null distribution. This test was implemented with the *sigclust* library in R ^28^ with number of simulations set to 10,000.

#### Longitudinal analyses

The *reval* modeling identifies subtypes from a single snapshot of early MSEL and VABS scores that represent LIMA features. To test whether the subtypes are different in later developmental trajectories in language, cognitive, motor, and adaptive domains, we ran longitudinal analyses on a subset of NDA individuals with repeated measures over time (Fig 1D). The reasoning behind conducting this analysis is that the subtypes within the A3D frameworks are predicted to have different developmental outcomes; while early developmental trajectories are meant to be suggestive of the rate of growth, they should also be predictive of future developmental outcomes. Longitudinal modeling was implemented using a linear mixed effect model (e.g., *lmer* from the *lme4* R library), whereby MSEL age-equivalent or VABS standard scores were the dependent variable and age, subtype and the age*subtype interaction were fixed effects and with random effects of age with random intercepts and slopes grouped by subject. We also ran longitudinal modeling with a second independent dataset from UCSD (n=1,487). Here we applied the NDA subtyping model to earliest intake (between 24-68 months) MSEL and VABS scores from the UCSD dataset to generate subtype labels (Fig. 1E). Similar linear mixed effect modeling was used to test age, subtype, and age*subtype interactions as fixed effects and with age as random intercepts and slopes group by subject.

#### Neuroimaging and gene expression data acquisition and preprocessing

All structural and functional neuroimaging data utilized in this work has been previously published in work examining ELO stratification (i.e. fMRI^17,18^, sMRI^19^). In this work, we take all data utilized in this past work and re-analyze it with different autism subtype labels (i.e. A3D Type I vs Type II). To briefly recap characteristics of this data and how it was initially collected, all neuroimaging data was collected during natural sleep at UCSD on a 1.5 Tesla General Electric MRI scanner. High-resolution T1-weighted IR-FSPGR (inversion recovery fast-spoiled prepared gradient recalled) anatomical scans were used for analyses of structural morphometry (surface area, cortical thickness) and for use during normalization steps in preprocessing fMRI data (parameters: TE (echo time) = 2.8 ms, TR (repetition time) = 6.5 ms, flip angle = 12°, bandwidth = 31.25 kHz, field of view = 24 cm, and slice thickness = 1.2 mm). Cortical morphometric measurements such as surface area (SA) and cortical thickness (CT) were derived from Freesurfer processing of the anatomical T1 data^19^. These morphometric measures were estimated from a genetically-sensitive parcellation known as GCLUST^29,30^. All SA and CT data analyzed here are identical to those reported in prior work^19^. fMRI data used in this work is identical to the data in previous papers on ELO^17,18^, and thus all details of the language paradigm and imaging acquisition parameters apply (i.e. single-echo echoplanar imaging (EPI) sequence; echo time = 30 ms; repetition time = 2500 ms; flip angle = 90°; bandwidth = 70 kHz; field of view = 25.6 cm; in-plane resolution = 4 × 4 mm; slice thickness = 4 mm; 31 slices). Preprocessed data was then input into single-subject general linear models (GLM) and was utilized for re-analysis^17^, but whereby new tests in this work specifically examine group differences using the new A3D stratification model (e.g., Type I vs Type II) and to compare the A3D stratification model to the previously reported ELO stratification model^17,18^. Both whole-brain activation mapping per each group and region of interest (ROI) analyses for between-group comparisons were used. In both whole-brain and ROI analysis models, sex and age at scan were included as covariates of no interest. Whole-brain mapping was thresholded using a voxel-wise FDR correction at q<0.05^31^. The ROIs utilized in this analysis were the same 4 meta-analytic ROIs used in prior work^17,18^, which were derived from Neurosynth for the term ‘language’ and comprise left and right hemisphere temporal and lateral frontal cortices. Model comparison between ELO and A3D models was implemented using the Akaike Information Criterion (AIC) statistic. Models with lower AIC are considered better and we considered the model sufficiently better if the difference in AICs was greater than 10^32^.

In this work, we also re-analyzed neuroimaging-gene expression relationships with partial least squares (PLS) analysis^33^, similar to previously reported in papers on ELO stratification^18,19^. The neuroimaging and blood leukocyte gene expression data re-analyzed was identical to those used in this past work^18,19^. However, in the current work we examine these relationships in the A3D stratification model (e.g., Type I vs Type II). To briefly recap how this data was initially collected and analyzed, all blood gene expression data was collected at UCSD. Four to six milliliters of blood was collected into EDTA-coated tubes from toddlers on visits when they had no fever, cold, flu, infections, or other illnesses, or use of medications for illnesses 72 h before the blood draw. Blood samples were passed over a LeukoLOCK filter (Ambion) to capture and stabilize leukocytes and immediately placed in a –20 °C freezer. Total RNA was extracted according to standard procedures and manufacturer’s instructions (Ambion). LeukoLOCK disks (Ambion cat no. 1933) were freed from RNAlater, and Tri-reagent (Ambion cat no. 9738) was used to flush out the captured lymphocytes and lyse the cells. RNA was subsequently precipitated with ethanol and purified though washing and cartridge-based steps. The quality of the mRNA samples was quantified according to the RNA Integrity Number (RIN): values of 7.0 or greater were considered acceptable^34^, and all processed RNA samples passed RIN quality control. Quantification of RNA was performed with a NanoDrop spectrophotometer (Thermo Scientific). Samples were prepared in 96-well plates at a concentration of 25 ng/μl.

RNA was assayed at Scripps Genomic Medicine for labeling, hybridization, and scanning with the Illumina BeadChip pipeline (Illumina) according to the manufacturer’s instructions. All arrays were scanned with the Illumina BeadArray Reader and read into Illumina GenomeStudio software (version 1.1.1). Raw data were exported from Illumina GenomeStudio, and data preprocessing was performed with the *lumi* package^35^ for R (http://www.R-project.org/) and Bioconductor (http://www.bioconductor.org/)^36^. Raw and normalized data are part of larger sets deposited in the Gene Expression Omnibus database (GSE42133 and GSE111175).

A larger primary dataset of blood leukocyte gene expression was available from 383 samples from 314 toddlers 1–4 years old. The samples were assayed with the Illumina microarray platform in three batches. The datasets were combined by matching the Illumina Probe ID and probe nucleotide sequences. The final set included a total of 20,194 gene probes. Quality-control analysis was performed to identify and remove 23 outlier samples from the dataset. Samples were marked as outliers if they showed low signal intensity (average signal two SD lower than the overall mean), deviant pairwise correlations, deviant cumulative distributions, deviant multidimensional scaling plots, or poor hierarchical clustering, as described elsewhere^37^. The high-quality dataset included 360 samples from 299 toddlers. High reproducibility was observed across technical replicates (mean Spearman correlation of 0.97 and median of 0.98). Thus, we randomly removed one of each of two technical replicates from the primary dataset. The 20,194 probes were then collapsed to ∼14,000 genes (14,313 for the fMRI-gene expression dataset^18^; 14,426 for the structural MRI-gene expression dataset^19^) on the basis of picking the probe with maximal mean expression across samples. Data were quantile normalized and then adjusted for batch effects, sex, and RIN. The batch-, sex-, and RIN-adjusted data were used in all further downstream analyses.

WGCNA analysis was used to reduce several thousands of highly correlated genes down to a less redundant number of gene co-expression modules to utilize in the PLS analysis. The WGCNA analysis is described in past papers^18,19^, and is briefly recapped here. WGCNA is implemented with the *WGCNA* library in R^38^. Correlation matrices estimated with the robust correlation measure of biweight midcorrelation and then converted into adjacency matrices that retained the sign of the correlation. These adjacency matrices were then raised to a soft power of 16. This soft power was chosen by finding the first soft power for which a measure of *R*2 scale-free topology-model fit saturated at least above *R*2 > 0.8^39^, and the slope was between – 1 and –2^40^. The soft-power-thresholded adjacency matrix was then converted into a topological overlap matrix (TOM) and then a TOM dissimilarity matrix (1 – TOM). The TOM dissimilarity matrix was then input into agglomerative hierarchical clustering with the average linkage method. Gene modules were defined from the resulting clustering tree, and branches were cut with a hybrid dynamic tree-cutting algorithm (deepSplit parameter = 4). Modules were merged at a cut height of 0.2, and the minimum module size was set to 100. Only genes with a module membership of *r* > 0.3 were retained within modules. For each gene module, a summary measure called the ME was computed as the first principal component of the scaled (standardized) module expression profiles, and these ME values were utilized in further PLS analyses. We also computed module membership for each gene and module. Module membership indicated the correlation between each gene and the ME. Genes that could not be clustered into any specific module were left within the M0 module, and this module was not considered in any further analyses.

Partial least squares (PLS) analysis^33^ was utilized to examine neuroimaging-gene expression relationships and was implemented in an identical manner as prior work^18,19^. To briefly recap, PLS allows for identification of latent imaging-gene expression variable pairs (LV) that maximally explained covariation in the dataset and which are uncorrelated with other latent variable pairs. The strength of such covariation was denoted by the singular value (*d*) for each brain–behavior LV, and hypothesis tests were made via with permutation tests on the singular values. Furthermore, identifying brain regions that most strongly contribute to each LV pair was made via bootstrapping, whereby a bootstrap ratio was created for each voxel and represented the reliability of that voxel for contributing strongly to the LV pattern identified. The bootstrap ratio was roughly equivalent to a *Z* statistic and could be used to threshold data to find voxels that reliably contribute to an LV pair. The PLS analyses reported here were implemented within the *plsgui* MATLAB toolbox (http://www.rotman-baycrest.on.ca/pls/). The imaging data for fMRI was single-subject t-statistic maps, while for structural MRI imaging data GCLUST parcellated surface area and cortical thickness measures were used. WGCNA module eigengene values were inserted for the gene expression data in the PLS analysis. For statistical inference on identified imaging-gene expression LV pairs, a permutation test was run with 10,000 permutations. To identify reliably contributing voxels/brain regions for imaging-gene expression LVs and to compute 95% CIs on correlations, bootstrapping was used with 10,000 resamples. Bootstrap ratios (BSR) were used to visualize patterns across the brain, with larger BSRs indicating stronger and more reliable contributions to significant imaging-gene expression LVs. The strength of imaging-gene expression correlations for significant LVs was displayed as a bar graph with 95% bootstrap CIs as error bars. Gene coexpression modules in which the 95% CIs did not encompass 0 were denoted ‘nonzero’ association modules. All other modules in which the 95% CIs included 0 were denoted ‘zero’ modules.

#### Gene set enrichment analyses

Our PLS analyses highlight non-zero modules that significantly contribute the imaging-gene expression association effects. Thus, we utilized genes from non-zero modules and then tested them for enrichment with an independent gene list of attenuated transcriptomic regional identity (ARI) genes, isolated in post-mortem cortical tissue of autistic individuals^41^. This analysis was done to test how genes isolated assayed from blood leukocytes and associated with imaging phenotypes in autism may overlap with genes isolated as dysregulated in their cortical patterning directly in cortical tissue of autistic patients. Gene set enrichment analyses were implemented using custom R code written by MVL that computes hypergeometric *P* values and enrichment ORs. The background pool for these enrichment tests set to the number of total genes assessed in our prior papers (e.g., 14,313^18^; 14,426^19^). After all enrichment tests were computed, results are interpreted only if the enrichment was statistically significant after FDR correction for multiple comparisons at a threshold of *q* < 0.05.

### Data availability

All data utilized in this work can be found in the National Institute of Mental Health Data Archive (NDA; https://nda.nih.gov). For the UCSD ACE data, the data are available on NDA through collection IDs 9, 2466, 2968, 2290 and 2115.

### Code availability

Analysis code to reproduce the analyses and figures is available on GitHub (https://github.com/IIT-LAND/a3d_msel_vabs). The *reval* Python library can be found on GitHub (https://github.com/IIT-LAND/reval_clustering) and the documentation at https://reval.readthedocs.io. The Shiny app that allows users to input their own MSEL and VABS data and get subtype predictions can be found at https://landiit.shinyapps.io/Autisms3D/.

## Results

### Detection of Type I versus Type II autisms in early development

The first objective of this work was to test whether it is possible to detect autism Type I and Type II subtypes in early development, as predicted by the A3D model. Stability-based relative clustering validation (*reval*)^16^ identified a two-cluster solution as the optimal solution and which generalizes with 98% accuracy in the independent NDA validation dataset (Fig 2A). SigClust analysis confirms that this solution is indeed indicative of true clusters that heavily deviate from the null hypothesis of that the data come from one originating multivariate Gaussian distribution (*p* = 9.99e-5; Fig 2B). The subtypes conform to the predicted Type I (‘disability’) versus Type II (‘difference’) distinction, as they show a marked separation between high versus very low scores across all LIMA features (Fig 2C). Notably, the distinction here is not based on hard cutoffs. Rather, distributions for all subscales/domains overlap to some extent, but effect sizes remain rather large throughout (Cohen’s d > 0.96) (Supplementary Table 1). The fact that distributions overlap and yet a classifier can still make accurate labelings with 98% accuracy indicates the robustness of such an approach over-and-above other heuristic methods typically used in clinical practice such as hard cut-off thresholding.

**Figure 2:**
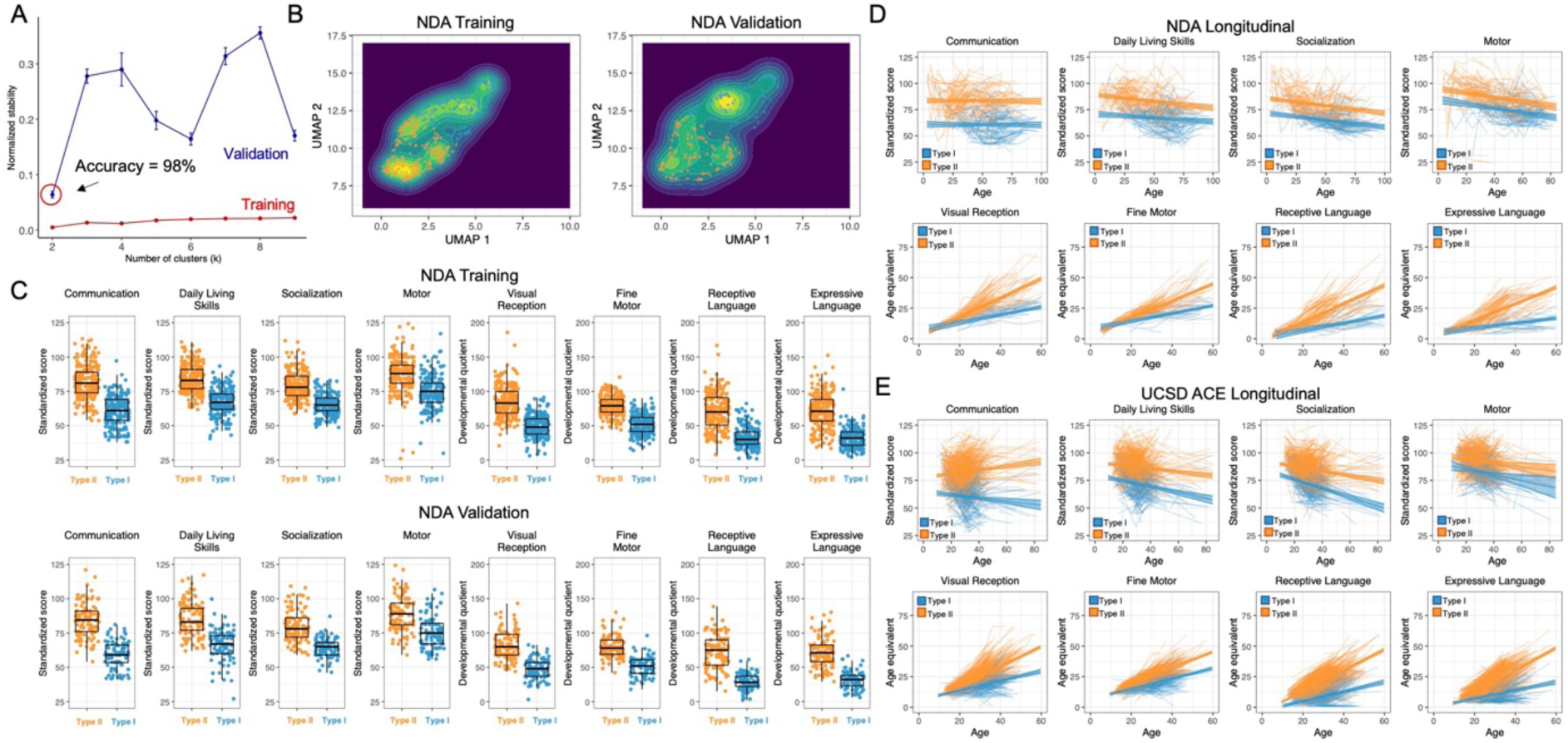
Robust and highly generalizable early distinctions between Type I versus Type II autisms. Panel A shows that a 2-subtype solution is the optimal clustering solution that minimizes normalized stability via stability-based relative clustering validation (reval). This solution generalizes with 98% accuracy in the held-out NDA validation dataset. Panel B shows UMAP density plots that depict separated peaks of distributions for Type I (blue) and Type II (orange) autism subtypes. Panel C plots the standardized scores across all MSEL and VABS features to describe how subtypes manifest as relatively low (Type I; blue) versus high (Type II; orange) scores across all features. Panels D-E show plots of developmental trajectories for the Type I (blue) and Type II (orange) autisms, respectively for NDA (D) and UCSD ACE (E) datasets.

### Differential developmental trajectories for Type I vs Type II autisms

We next tested if A3D Type I versus Type II autisms differed in terms of subsequent developmental trajectories taken over the first decade of life (Fig 2D-E). For each MSEL subscale, we found highly significant age*subtype interactions in both NDA and UCSD ACE datasets (Supplementary Tables 2-3). This interaction effect can be described by slower and less steep trajectories for Type I compared to Type II and indicates that Type II autism develops language, fine motor, and non-verbal cognitive skills at a much faster rate. For VABS trajectories we did not find any significant age*subtype interactions in the NDA, although the slopes appear qualitatively different (Supplementary Table 2). In contrast, significant age*subtype interactions appear in the UCSD ACE dataset for all VABS domains except for the motor domain (Supplementary Table 3). Notably, the larger longitudinal sample size for the UCSD ACE dataset (n =1,487) may imply that such subtle yet significant effects here were not detected in NDA because of lower statistical power. VABS trajectory differences can be described as Type II autism have much less steep decline in skills over time compared to Type I autism. Overall, these results showcase that Type I and Type II autisms can be characterized by not just initial large differences in early LIMA features, but also by differential rates of development over the first decade of life. Thus, if Type I vs Type II autisms are a distinction between profound disability versus difference, these results show that those descriptions are increasingly more prominent as development progresses.

### Differential functional neural responses to speech

In the next analysis we examined fMRI responses to speech between A3D Type I and Type II autisms. Prior work using early language outcome (ELO) subtype labels showed that autism with poor ELO does not robustly activate language-sensitive superior temporal cortices^17,18^. Since the same fMRI data was re-analyzed in this work, we first examined how well the A3D labels might correspond to the ELO subtype labels. In contrast to the ELO model, the A3D model breaks down into a somewhat different distinction (*χ*^2^ = 14.21, *p* = 0.0001) (Supplementary Table 4). Type I individuals in A3D are about 17% of all autistic individuals in the fMRI dataset, and all Type I individuals in A3D would be labeled in the Poor subtype of the ELO model. In contrast, all individuals labeled in the Good subtype under the ELO model fell into the Type II A3D subtype (Supplementary Table 4). This contrast between models and how labels are made indicates that the Type I label from A3D is much more conservative than the ELO model.

Despite the considerable difference in subtype labels between the models, whole-brain maps show a quite similar activation patterns in each subtype between ELO and A3D models. The robust activation patterns observed in TD and LD/DD non-autistic comparison groups are largely preserved, particularly in left hemisphere superior temporal cortex, in the ELO Good and A3D Type II subtypes. In contrast, very little is identified in the ELO Poor and A3D Type I subtypes (Fig 3A). This qualitatively may suggest that very similar fMRI differences are apparent across the ELO and A3D models. To test this in a more targeted way for specific brain regions sensitive to language, we conducted region of interest (ROI) analyses around 4 regions (bilateral temporal and frontal cortex regions) that meta-analytic evidence shows are sensitive to language processing^17,18^. Here we find robust evidence that the ELO pattern of the Poor subtype showing a lack of left hemisphere superior temporal cortex (LH temporal) response to speech is similarly evident in the Type I subtype from A3D, but with somewhat larger effect sizes in A3D (Fig 3B; Supplementary Table 5A). Model comparisons with the Akaike Information Criterion (AIC) show that ELO and A3D models are equally good at explaining variance in functional neural responses to speech (*ELO AIC* = −446.59; *A3D AIC* = −447.92, Supplementary Table 5B). The slightly lower AIC for the A3D model may be due to marginally stronger effect sizes for TD vs Type I and LD/DD vs Type I, compared to the ELO TD vs Poor and LD/DD vs Poor comparisons (Fig 3B). Similar effects also emerge with ELO and A3D models showing very similar AIC values in RH superior temporal cortex (RH temporal) (Fig 3C; Supplementary Table 5B, while little to no group differences are apparent in LH and RH frontal ROIs for either ELO or A3D (Supplementary Table 5B).

**Figure 3:**
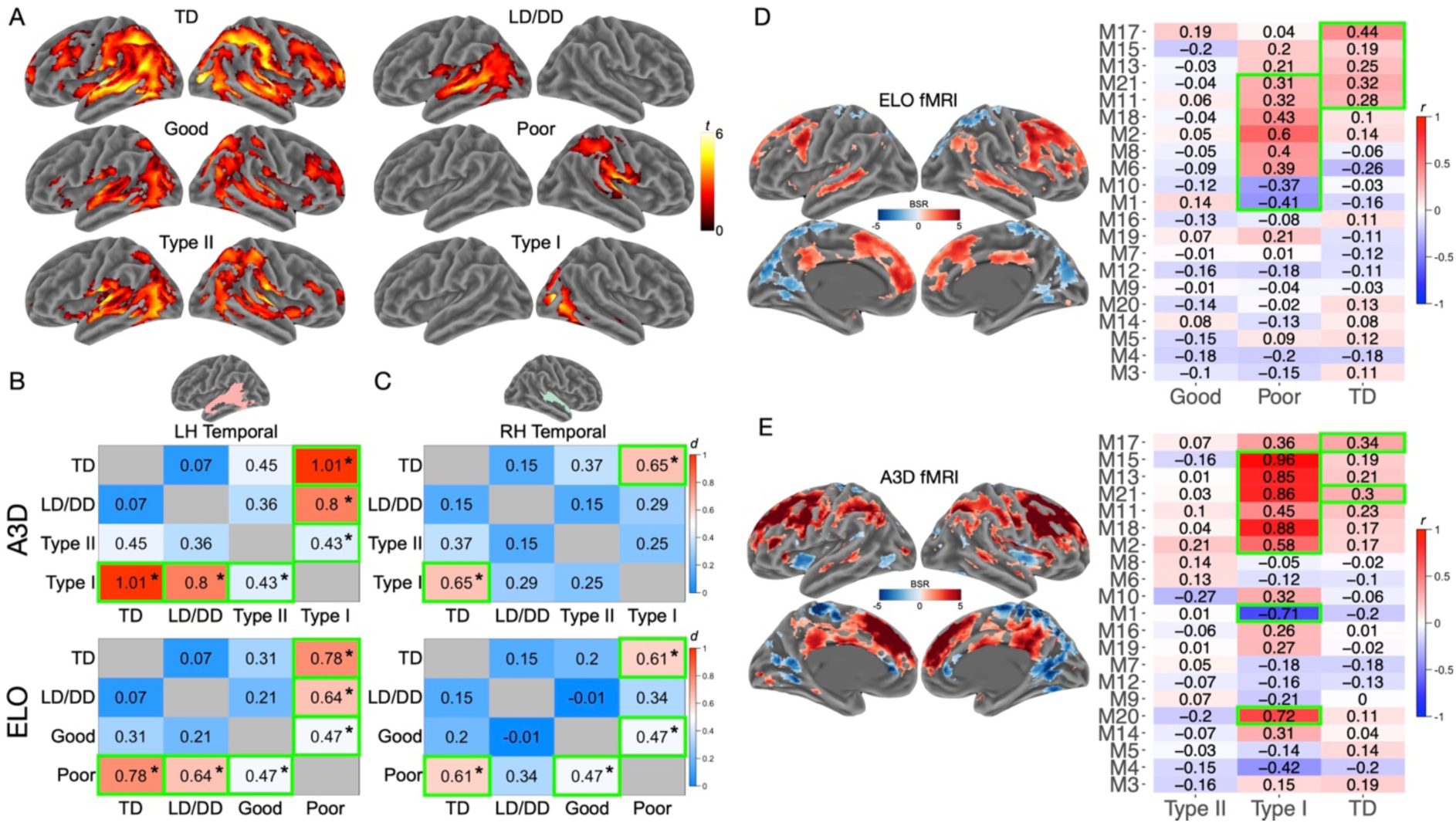
Functional neural response to speech and associations with gene expression. Panel A depicts whole-brain analysis maps for each of the groups under study. All maps are thresholded at voxel-FDR q<0.05, except for LD/DD, Poor and Type I groups, which are thresholded at cluster-forming p<0.05 and cluster FDR q<0.05 for visualization purposes. Panel B depicts standardized effect sizes (Cohen’s d) for the LH temporal ROI for all pairwise group comparisons for A3D (top) and ELO (bottom) stratification models. The asterisks and green outlined cells indicate statistical significance at p<0.05. Similar effect size heatmaps are shown in panel C for the RH temporal ROI. Panels D and E show results for the PLS analysis of fMRI-gene expression relationships in the ELO (D) and A3D (E) models. The whole-brain map show brain bootstrap ratios (BSR) to indicate which voxels most strongly contribute to the fMRI-gene expression relationship. The heatmaps to the right on each panel shows each gene co-expression module (rows) and groups (columns). The modules of importance are highlighted with green outlines (non-zero modules) as they contain a correlation whose 95% bootstrap CIs do not encompass zero.

### Differential large-scale associations between fMRI response and gene expression

We next compared how functional neural response to speech is related to gene expression patterns measured in blood. In our prior work on the ELO model, we found evidence for a large-scale fMRI-gene expression relationship whereby such relationships are relatively distinct between TD and autism subtypes^18^ (Fig 3D). The same analysis applied to the A3D model reveals a very similar relationship (LV1: *d* = 110.73, *p* = 1.99e-4, *percentage covariance explained* = 34.16%), comprising many of the same frontal and temporal regions, but additionally showing prominent contributions of lateral parietal areas connected by the arcuate fasciculus in language-sensitive areas^42^ (Fig 3E). Additionally, the correlations between specific non-zero gene co-expression modules apparent in the Type I subtype (Fig 3E) are much stronger than similar relationships observed in the ELO Poor subtype (Fig 3D). Thus, much like the prior analysis showing similar differences in fMRI activation response to speech, associations with functional genomic mechanisms also appear to be quite similar in A3D vs ELO, albeit with slightly stronger effect in A3D.

### Differential genomic cortical patterning of cortical surface area

In prior work we also reported that genomic cortical patterning of surface area and cortical thickness manifest differently in ELO subtypes^19^. In the next analysis, we revisit that same dataset and apply the A3D model. Similar to the comparison of fMRI ELO vs A3D models in terms of sample sizes, in this analysis of structural imaging phenotypes the A3D model was again more conservative in labeling individuals as Type I (17% of all autistic individuals) compared to the ELO model Good vs Poor distinction (*χ^2^* = 13.36, *p* = 0.0002) (Supplementary Table 4). PLS analyses of surface area in the ELO model revealed on one significant LV pair, interpreted as a normative cortical patterning effect with similar associations found in the TD and autism Good subtype, whereas such patterns were absent in the autism Poor ELO subtype^19^ (Fig 4A). In contrast, analysis of the A3D model reveal 2 significant LV pairs, with LV1 being an effect showing effects exclusively in the Type I subtype (LV1: *d* = 4.22, *p* = 1.99e-4, *percentage covariance explained* = 26.78%; Fig 4B). The LV2 recapitulates the normative surface area patterning effect found in the ELO model (LV2: *d* = 3.57, *p* = 1.99e-4, *percentage covariance explained* = 19.21%; Fig 4C), whereby similar correlations are found in TD and Type II. However, LV2 also reveals opposite effects occurring for Type I in some modules (e.g., M4, M6), and additional modules with important non-zero effects that are exclusive to Type I (e.g., M11, M12). Thus, although retaining a similar normative surface area patterning effect, the A3D model is additionally sensitive to an effect for how surface is atypically patterned specific to the Type I subtype.

**Figure 4:**
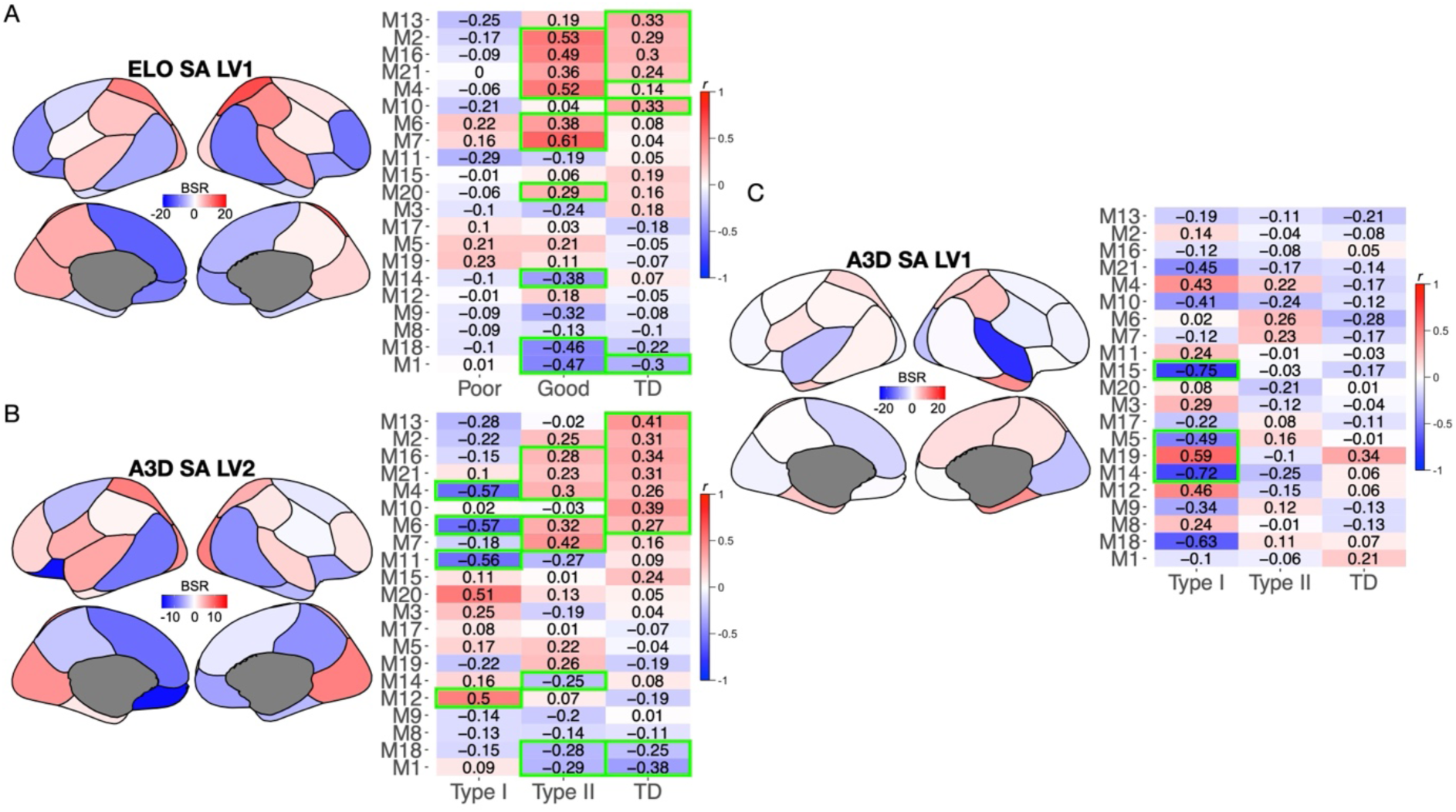
Gene expression associations with cortical surface area. Results for PLS analyses on the phenotype of cortical surface area for the ELO (A) or A3D (B, A3D SA LV2; C, A3D SA LV1) models. The parcellated brain maps show brain bootstrap ratios (BSR), whereby more extreme BSRs indicate brain regions that more strongly contribute to the LV relationship. The heatmap on the right of each panel shows correlations with each gene co-expression module (rows) and each group (columns). The important non-zero modules are highlighted as cells with green outlines.

### Differential genomic cortical patterning of cortical thickness

Next, we looked at genomic patterning of cortical thickness. The ELO model had previously revealed 2 different effects on cortical thickness – LV1 being a normative effect with similar associations in TD and the autism Good ELO subtype (Fig 5A) and LV2 being an atypical effect on cortical thickness specific to the autism Poor ELO subtype^19^ (Fig 5B). The A3D model reveals only one significant LV pair (A3D CT LV1: *d* = 4.98, *p* = 9.99e-5, *percentage covariance explained* = 35.33%; Fig 5C). However, A3D CT LV1 is an effect that encompasses both the normative effect (e.g., similar non-zero modules (M1, M16) shared by TD and Type II) and an atypical effect specific to Type I (e.g., non-zero modules that are unique to Type I (M3, M5, M6, M9, M12), or which appear in Type I but in the opposite directionality (M7) from TD and Type II). Qualitatively, similar non-zero modules appear present for both TD and Good or Type II subtypes in both models. In contrast, somewhat different non-zero modules appear that are unique to the Poor group in ELO CT LV2 compared to Type I in A3D CT LV1.

**Figure 5:**
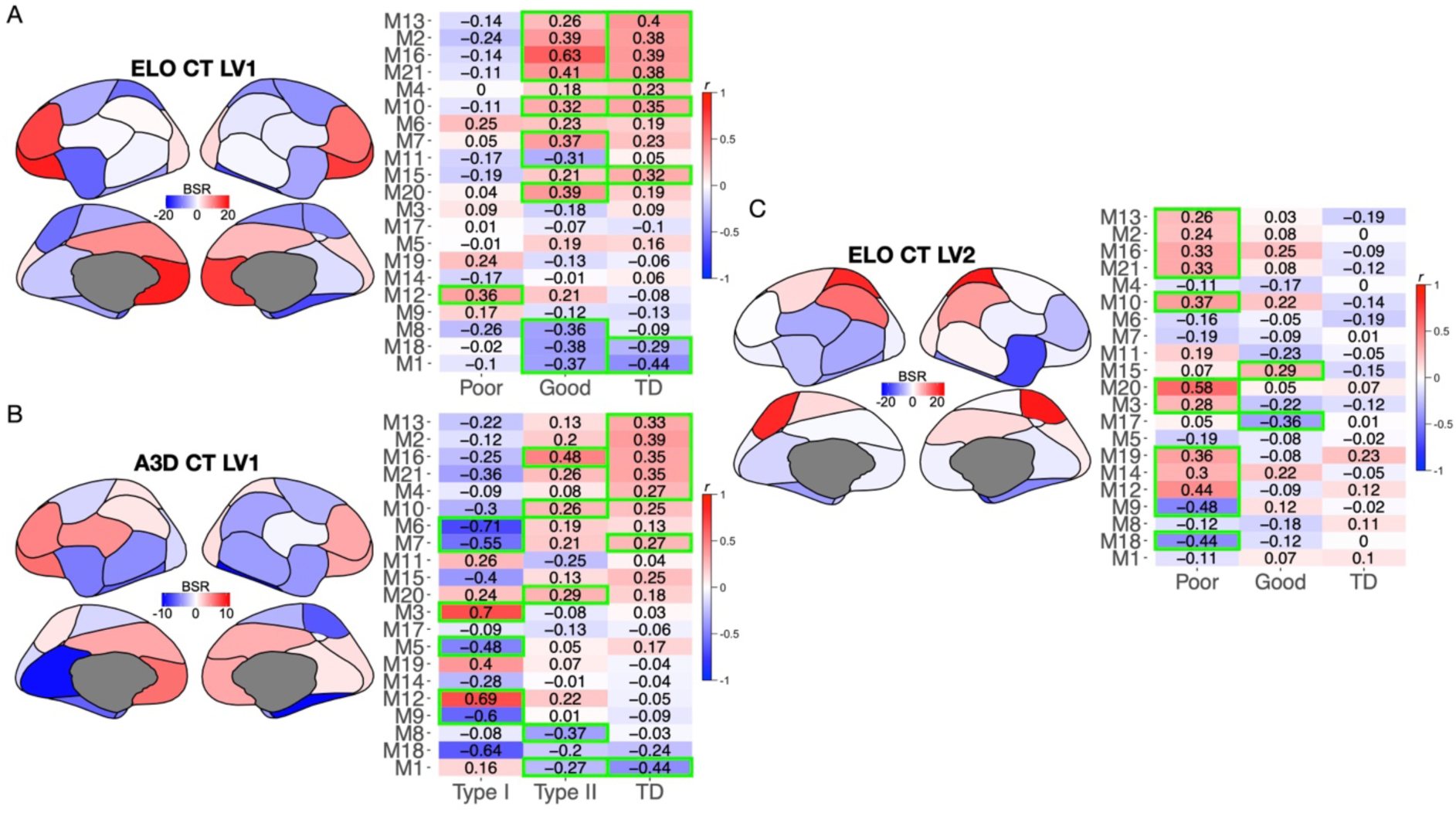
Gene expression associations with cortical thickness. Results for PLS analyses on the phenotype of cortical thickness for the ELO (A, ELO CT LV1; C, ELO CT LV2) or A3D (B, A3D CT LV1) models. The parcellated brain maps show brain bootstrap ratios (BSR), whereby more extreme BSRs indicate brain regions that more strongly contribute to the LV relationship. The heatmap on the right of each panel shows correlations with each gene co-expression module (rows) and each group (columns). The important non-zero modules are highlighted as cells with green outlines.

### Overlap with genomic cortical patterning effects in post-mortem cortical tissue

A caveat to the analyses showing large-scale associations (e.g., cortical patterning effects) between imaging phenotypes and gene expression is that these data deal with gene expression patterns measured peripherally in blood leukocytes. However, a recent paper on broad transcriptomic dysregulation across cortical regions in post-mortem cortical tissue reveals a potentially similar kind of pattern effect for how gene expression is dysregulated across the cortex in autism^41^. Thus, we took the opportunity here to assess whether there is indeed overlap between genes that show an atypical transcriptomic regional identity (ARI) effect in post-mortem cortical tissue and the genes that pop up in our imaging-gene expression association analysis (i.e. genes from non-zero modules). Here we find that fMRI non-zero module genes from both the ELO and A3D model significantly overlap with ARI genes identified in post-mortem cortical tissue^41^ (Fig 6A). With regard to structural cortical patterning of surface area and cortical thickness, we also find significant overlap between Gandal’s ARI gene set and genes from PLS effects that are interpreted as showing normative patterning effects in TD and ELO Good or A3D Type II subtypes (e.g., similar non-zero modules and associations). In contrast, PLS LVs interpreted as specific atypical effects in ELO Poor or A3D Type I subtypes do not show strong overlap with Gandal’s ARI gene set (Fig 6B; Supplementary Table 6). The A3D CT LV1 effect, characterized by a mixture of shared normative effects between TD and Type II versus specific atypical effects in Type I, was an exception, as the enrichment with ARI genes was trending on significance after FDR correction (*FDR q* = 0.055; Supplementary Table 6). Thus, these analyses are primary validation for the idea that imaging-blood gene expression associations revealed here and in our prior work can pick up on similar genes that when measured in cortical tissue, show atypical gene expression patterning dysregulation in autism. In particular, when our analyses of blood gene expression identify normative patterning effects shared in TD and ELO Good or A3D Type II subtypes, these effects are quite similar to those appearing in the ARI gene list from post-mortem cortical tissue.

**Figure 6:**
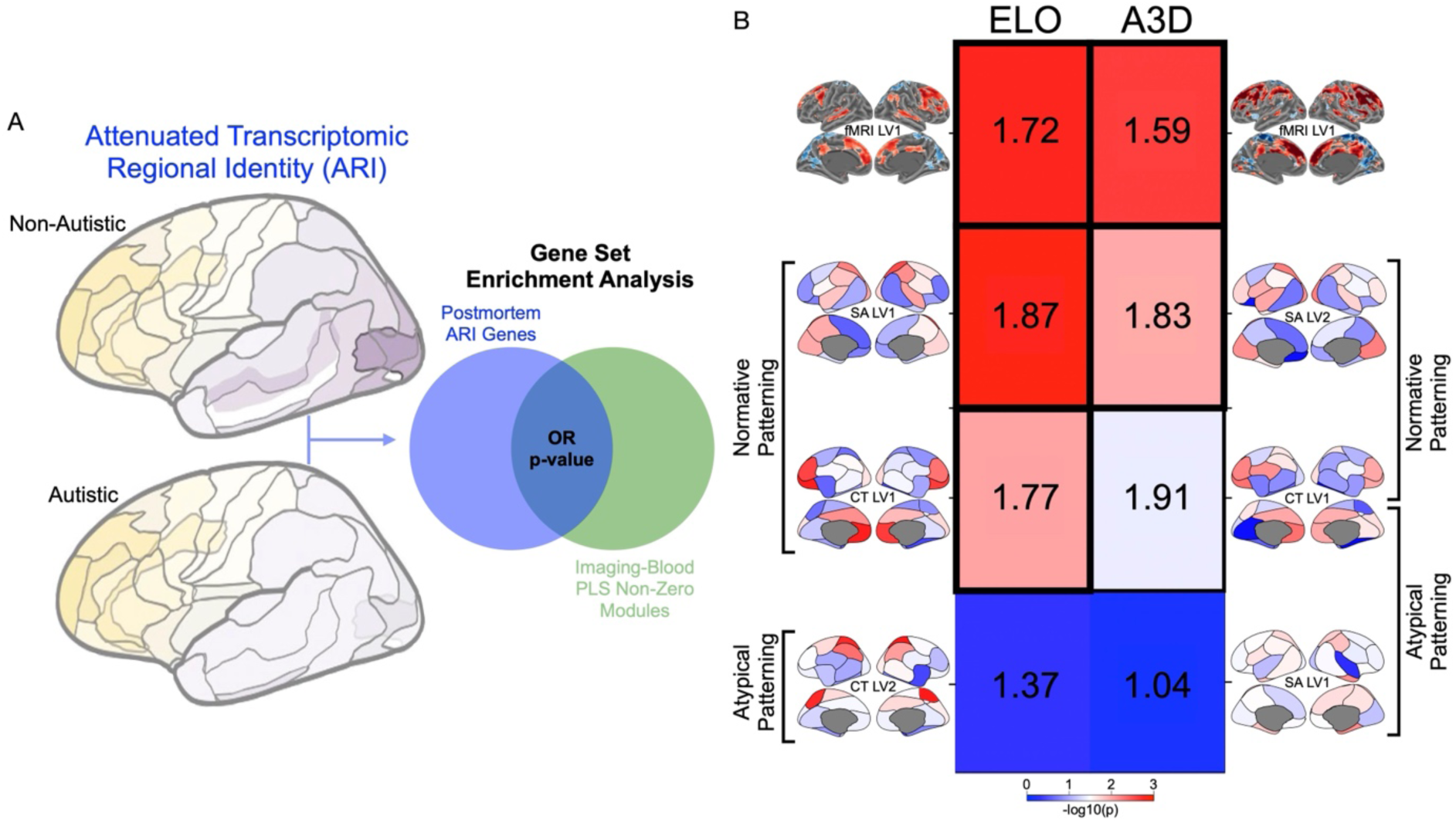
Enrichment analysis showing overlap between attenuated transcriptomic regional identity (ARI) genes detected in post-mortem cortical tissue and non-zero modules detected in imaging-gene expression PLS analyses. Panel A depicts a schematic of how these enrichment analyses were conducted. A gene set known as attenuated transcriptomic regional identity (ARI) genes were extracted from a prior study on post-mortem cortical tissue in autistic patients^41^. These ARI genes are genes that have substantial between-region gene expression differences in non-autistic brains, but much more attenuated regional identity differences in gene expression in autism. ARI genes are depicted in blue in the Venn diagram. In green, the Venn diagram shows our set of genes isolated from PLS imaging-gene expression association analyses. The degree of overlap is then tested with enrichment odds ratios and hypergeometric p-values. Panel B shows a heatmap of the enrichment odds ratios (numbers in the center of each cell) whereby color in each cell indicates the −log10(p-value) for each hypergeometric test. Cells with a thick black outline indicate enrichment tests that pass at FDR q<0.05, while cells with smaller thin black outline pass at FDR q<0.1. The columns in the heatmap represent comparisons when the gene list derives from the ELO (left) or A3D (right) model. Each row of the heatmap indicates a different gene list extracted from PLS analyses, with the top row indicating genes from non-zero modules in PLS analyses that examine fMRI responses to speech. The remaining rows depict comparisons where the PLS gene lists come from associations with surface area (SA) or cortical thickness (CT) and we have annotated with rows can be interpreted as effects indicative of ‘normative patterning’ effects (effects driven by TD correlations and which are shared in Good ELO or Type II A3D subtypes) versus atypical patterning effects (e.g., effects driven specifically in the ELO Poor or Type I A3D subtypes.

## Discussion

This work offers a key proof-of-concept for the A3D model approach to stratifying autism into phenotypically, biologically, and developmentally-meaningful subtypes. The A3D model suggests that a first-level split in the autism population should be between subtypes differentially characterized by profound disability (Type I) versus difference (Type II) and that such a distinction is evident from early in life and continually emerges throughout later development. At the basis for differentiating Type I versus Type II autisms under A3D are the non-core LIMA features. An unsupervised data-driven modeling approach can identify these types in an automated fashion with high degree of accuracy in generalizing to new data. To accelerate research progress in the field testing these subtypes, we have developed a freely available web application that will allow researchers to insert their own MSEL and VABS data and immediately get subtype labels (https://landiit.shinyapps.io/Autisms3D/).

In addition to providing a robust, reproducible, and highly generalizable prediction model for detecting Type I versus Type II autisms, we have also provided replicable evidence in two independent longitudinal datasets demonstrating that identification of such subtypes in early points in development is predictive of later differences in developmental trajectories over the first decade of life. This key feature of the results brings together the ‘developmental’ component of A3D by showing that the subtypes can be identified early and that such early generated subtype labels are indicative of different developmental paths as they grow older.

In this work, we also tested the A3D model to determine whether the subtype labels are sensitive to differences at the level of neuroimaging phenotypes and relationships to gene expression. The A3D model highlights several distinctions that are very similar in nature to the previous ELO stratification model^17–19^. The similar biological sensitivity of A3D and ELO is important, since the two models do differ substantially in terms of labels, with A3D potentially being much more conservative. Some subtle and qualitative differences did emerge between A3D and ELO with respect to how neuroimaging phenotypes are related to gene expression. First, regarding fMRI-gene expression associations, it was clear that the A3D model generally produces associations in Type I that are a bit stronger than those seen in the ELO Poor subtype. Second, regarding surface area-gene expression associations, A3D revealed two kinds of associations – one being a normative effect shared between TD and Type II, and a separate atypical effect apparent only in Type I. This can be contrasted to only a normative genomic patterning effect on surface area in the ELO model. Finally, whereas CT-gene expression associations in the ELO model comprised two orthogonal effects – one normative and shared between TD and Good subtype, while another is atypical specifically in the Poor subtype – the A3D model identifies only one latent variable pair that captures both the CT normative patterning effect in TD and Type II, but also capture atypical patterning in different co-expression modules for Type I. These types of subtle and qualitative differences may be important for distinguishing A3D from the ELO model. Given that A3D tends to identify somewhat larger effects alongside unique imaging-gene expression relationships for Type I and/or relationships with opposing directionality in Type I compared to Type II and TD, it could be inferred that A3D may be better than ELO at highlighting biology in the most profoundly affected autistic individuals compared to the ELO model. However, we also emphasize the large degree of similarities between results from A3D and ELO. This overlap illustrates that both ELO and A3D are detecting differential biology that is otherwise hidden from case-control modeling. A limitation/caveat to underscore about the neuroimaging results is that current sample sizes, particular for Type I in A3D, are relatively small. Thus, future work should attempt to replicate and extend these findings in larger samples.

To conclude, we demonstrate that the autism Type I and Type II subtypes, as described by A3D theoretical framework, are present in early development and that their early developmental trajectories are markedly different in language, motor, intellectual and adaptive skills. This work proves the usefulness of the A3D model in parsing autism heterogeneity highlighting important differences that, already proved to be in the short term, have the potential to be a good prognostic model for long term outcomes. To provide a complete tool for A3D model application, future works should aim to implement a stratification model to distinguish Type I and Type II subtypes in autistic individuals older than 68 months.

## Acknowledgments

This project was supported by funding from the European Research Council (ERC) under the European Union’s Horizon 2020 research and innovation programme under grant agreement No 755816 (AUTISMS) (ERC Starting Grant to MVL) and under the European Union’s Horizon Europe research and innovation programme under grant agreement No 101087263 (AUTISMS-3D) (ERC Consolidator Grant to MVL). Neuroimaging data and longitudinal phenotypic data from UCSD ACE was collected with the support of the following grants: NIMH R01-MH080134 (KP), NIMH R01-MH104446 (KP), NFAR grant (KP), NIMH Autism Center of Excellence grant P50-MH081755 (EC, KP), NIMH R01-MH036840 (EC), NIMH R01-MH110558 (EC), NIMH U01-MH108898 (EC), NIDCD R01-DC016385 (EC, KP, LE, MVL), CDMRP AR130409 (EC), and the Simons Foundation 176540 (EC).

## Author Contributions

Conceptualization: MVL. Methodology: MVL, VM, IS. Formal analysis: MVL, VM, IS. Investigation: MVL, VM, IS, LE, KP, EC. Writing - original draft preparation: MVL, VM, IS. Writing - review and editing: MVL, VM, IS, LE, KP, EC. Visualization: MVL, VM, IS. Supervision: MVL. Project administration: MVL. Funding acquisition: MVL.

## Competing Interests

All authors have no competing interests to declare.

## upplementary Table Legends

**Supplementary Table 1: Standardized effect sizes describing magnitude of difference between MSEL and VABS scores for Type I versus Type II in NDA.** Magnitude of difference between Type I versus Type II are reported in this table as standardized effect sizes (Cohen’s d) for all MSEL and VABS scales in NDA Training and Validation sets.

**Supplementary Table 2: Linear mixed effect models (LMEs) for longitudinal VABS and MSEL data from the NDA dataset.** LMEs are computed with a fixed effect of age, subtype, and age* subtype interaction. Random effects are intercepts and slopes grouped by subjects.

**Supplementary Table 3: Linear mixed effect models (LMEs) for longitudinal VABS and MSEL data from the UCSD ACE dataset**. LMEs are computed with a fixed effect of age, subtype, and age* subtype interaction. Random effects are intercepts and slopes grouped by subjects.

**Supplementary Table 4:** Sample sizes for fMRI and MRI datasets, broken down by ELO and A3D subtype labels. Confusion matrices are shown in these tables to showing correspondence between A3D and ELO autism subtype labels in the UCSD ACE fMRI and MRI datasets.

**Supplementary Table 5: AUTISMS-3D model ROI analysis.** Table A shows the results from the ROI analysis of the AUTISMS-3D model; ANOVA’s F and p values are reported for the effects of Subtype, Sex, and Age at scan. Significant p-values (<0.05) are highlighted. Table B reports the AIC comparison between the AUTISMS-3D and ELO models. Delta AIC are reported as absolute values, the best model is highlighted.

**Supplementary Table 6. Gene set enrichment analysis.** This table shows results for the gene set enrichment analysis between Partial Least Square (PLS) gene list and attenuated transcriptomic regional identity (ARI) genes. Each row indicates a different gene list extracted from PLS analysis. LV1 = Latent Variable 1; SA = Surface Area; CT = Cortical Thickness; OR = Odds Ratio. Normative refers to patterning effect driven by TD and Type II/ELO Good individuals, while Atypical refers to patterning effects driven by Type I/ELO poor.

## Notes

### Competing Interest Statement

The authors have declared no competing interest.

### Author Declarations

This study was approved by the Institutional Review Board at University of California, San Diego.

